# A Simple Covid-19 Epidemic Model and Containment Policy in France

**DOI:** 10.1101/2020.04.25.20079434

**Authors:** J.-P. Quadrat

## Abstract

We show that the standard SIR model is not effective to predict the 2019-20 coronavirus pandemic propagation. We propose a new model where the logarithm of the detected population number follows a linear dynamical system. We estimate the parameters of this system and compare models obtained with data observed from different countries. Based on the given estimator and results obtained with the Pr. Raoult’s treatment, we affirm with a reasonable degree of confidence that his “test-treat-noconfine” policy was less expensive in human lives than the”confine and wait for a proved treatment” policy adopted by the French government.

## 1 Introduction

The forecast of the spread of the Covid epidemic is very important to take governmental decisions such as containment policy. In France, the decision of the population containment has been taken on the basis of the possible risk of 500 000 deaths forecasted by the Ferguson model (a student of Ferguson belonged to the scientific committee dedicated to Covid epidemic). At the same committee, Pr. Raoult’s recommendation was not to confine the population but to test, confine and treat infected people. In the end, the political decision has been the containment given the dramatic risk of 500 000 deaths. In our opinion, Pr. Raoult’s treatment was the correct one, able to reduce the mortality to a regular influenza epidemic.

The Ferguson’s model [3] is a renewal stochastic model using the SIR (Susceptible-Infected-Recovered) with a time-varying reproduction number that predicts the average number of secondary infections at a given time. Ferguson said : “The renewal model is related to the Susceptible-Infected-Recovered model, except the renewal is not expressed in differential form”. Here, after having shown that SIR deterministic dynamical models are not effective to forecast the number of infected-detected people, we will propose a new model. It is a dynamical linear model forecasting the logarithm of the cumulated infected-detected population. A first-order linear time-invariant model fits well with the observations. Explicit formulas can be given which are quite good at least when the policy test to detect infected people or the containment policy does not change too much.

## 2 SIR models are not effective to forecast the epidemic evolution

The SIR (Susceptible-Infected-Recovered) model distinguishes three kind of people in a given population. We can denote by *S* the proportion of the susceptible population to be infected, *I* the proportion able to infect and by *R* the proportion which has recovered and which is not able to transmit the infection. We have *S* + *I* + *R* = 1. This model assumes that the number of newly infected people is proportional to the product of the number of infected by the number of susceptible ones. Based on this assumption the following differential equations describing the evolution of the three variables are proposed:

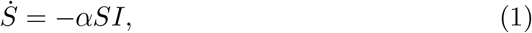

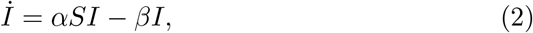

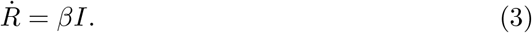

Where *α* can be seen as a reproduction number by unit of time and *β* is a recovering factor by unit of time.

We think that this standard SIR model must be improved a little bit. It says nothing on the immune population at the end of the epidemic on the contrary to the following discrete-time model:

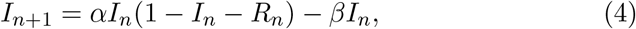

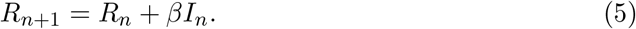

In equilibrium we have *R* = lim_*n*_ *R*_*n*_ and *I* = lim_*n*_ *I*_*n*_, then *R* satisfies 1 = *α*(1 *− R*) *− β* which gives *R* = 1 *−* (1 + *β*)*/α*. This is the well-known formula when *β* = 0 for the proportion of the infected people at the end of the epidemic. The continuous-time version of this model gives the following differential equations:

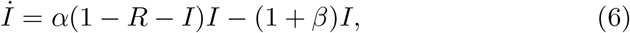

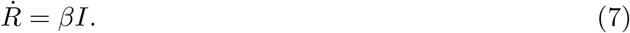

Using the new variable *T* = *I* + *R* instead of *I*, the system can be written :

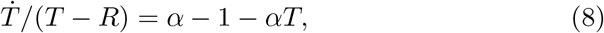

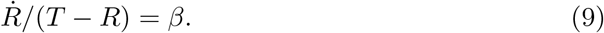

This form is useful to estimate parameters of *α* and *β*. The variable *T* will be called infected-recovered people^1^.

In more sophisticated models, we can distinguish the infected in classes according to the time since they have been infected. In other cases, we cut out the space into zones. In stochastic models, after a stochastic random time, a stochastic reproduction appears. Sometimes it is supposed that the *α* and *β* coefficients depend on time. But the main point is bilinearity : the new infected are proportional to the product of susceptible and infected people. The collective immunity corresponds to the fact that the recovered people cannot become able to contaminate.

For a given epidemic, to be able to forecast the evolution, we have to estimate the two numbers *α* and *β*. The problem is that we are not able to observe any of the two variables *T, R*. We can test some people or observe some symptoms but we cannot observe precisely the number of infected people.

At the beginning of the infection, as nobody has recovered, we can suppose that *R* = 0. If we suppose that the number of observed infected-recovered people is proportional to the infected-recovered ones (*y* = *NT* where *N* is an unknown parameter and *y* denotes the observed infected-recovered cases) we obtain the evolution equation of *y* :

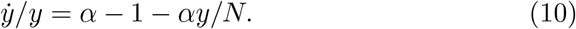

This equation can be written:

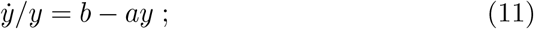

with *b* = *α −* 1 and *a* = *α/N* which can be easily estimated from the observation *y* by a simple linear regression.

Let us see the results on the French data for the covid epidemic. The Figure-1 gives the observation in green of 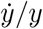, the regression *b − ay*(*t*) in black as function of time. We observe that the result is very bad. A better regression is easily obtained noticing that *y*(*t*) grows exponentially and therefore that log(*y*(*t*)) is almost proportional to time. The blue line gives the corresponding regression *B − A* log(*y*(*t*)).

**Figure 1:**
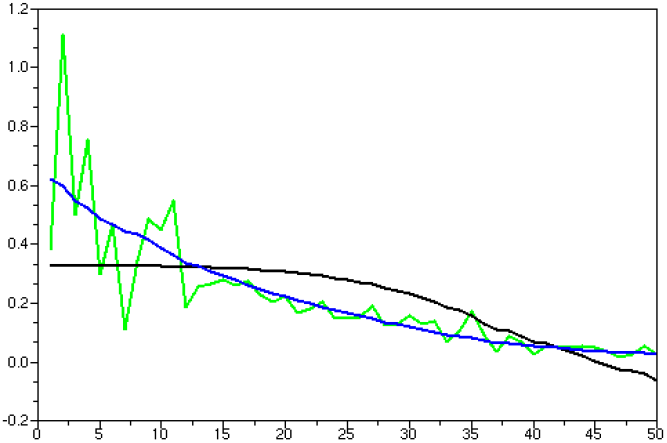
Green : observed 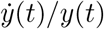. Black: *b−ay*(*t*). Blue: *B−A* log(*y*(*t*)).

Therefore a better model than the SIR model is given by :

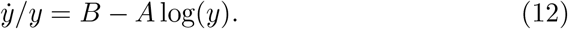

If we see the term “*−A* log(*y*)” as a brake on the exponential epidemic growing. This brake cannot be explained in immunology terms as the standard brake of the SIR model “*−ay*”. It is not clear to understand its origin. A possible explanation is a sociological reaction of the population. During an epidemic, people react to avoid being infected and this reaction would be proportional as the log of the affected population.

Using the new variable *z* = log(*y*) we obtain the simple model:

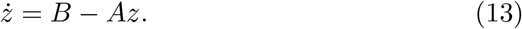

Therefore we have:

The main point of the paper.

The logarithm of the observed number of people infected during the covid epidemic denoted by *z*(*t*) can be well approximated by a first-order time invariant stable linear dynamical system *ż* = *−Az*+*B* and therefore the observed number of infected people *y*(*t*) can be parameterized by the three numbers *A, B, T* with the explicit formula:

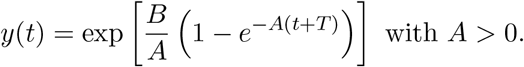

In [2] Bhardway proposed a linear dynamical model:

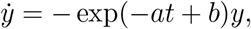

where he fitted *a* and *b* from observations. The main difference is that this model is time-variant and the fit is done according to time. Our model is time-invariant and the fit is according to the logarithm of the observation. The logarithm of the observation is not completely linear with respect to time. We can observe this curvature in Figure-1. Observations in green do not follow exactly a straight line but a little curved one. The blue line is better than any straight line. In [2] a fit with an exponential time is done. In the end, the two points of view give the same parameterization.

In Figure-2a we compare, in the French case, the cumulated numbers of observed positive people given by our model in blue with the SIR ones (when *β* = 0) in black and the observed ones in green.

**Figure 2:**
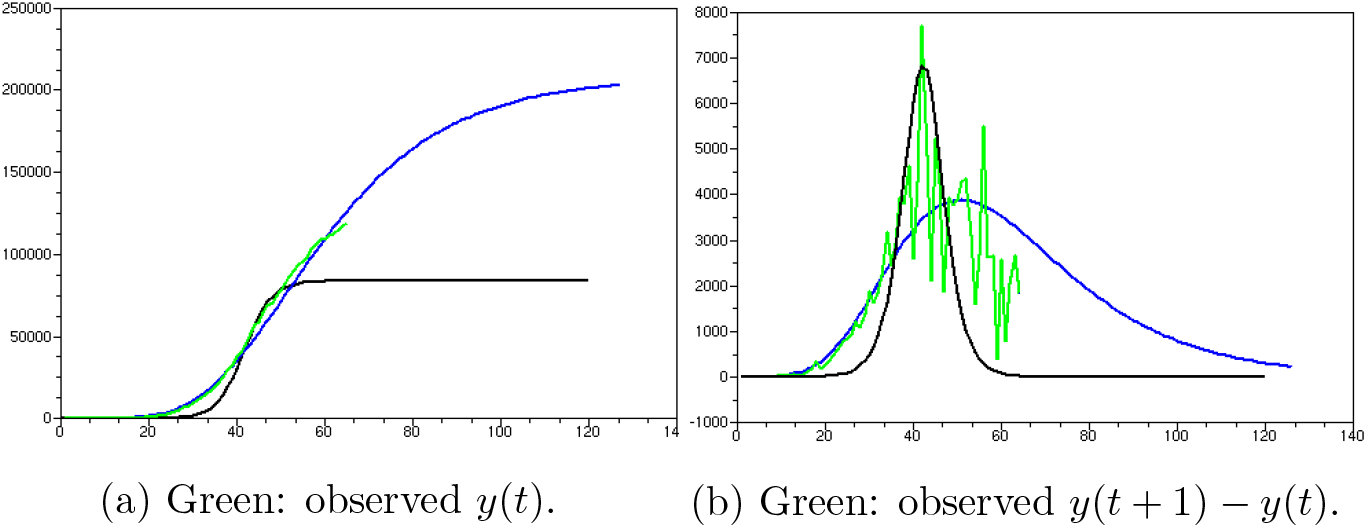
Black: SIR estimation. Blue: our estimation.

In Figure-2b we compare, in the French case, the daily numbers of observed positive people given by our model in blue with the SIR ones (when *β* = 0) in black and the observed ones in green. The estimated parameters for the SIR model are *α* = 1.327, *N* = 340 000, *β* = 0.02. In the simulation we have neglected *β* (*β* = 0). This comparison shows that it is difficult to have confidence in the SIR model to forecast the evolution of the covid epidemic.

## 3 Forecast for some countries

Before using the proposed model to forecast it is useful to have an idea of the robustness of the prevision. The prediction of the logarithm of the cumulated infected-detected people number is in general very good. But a small errors on the logarithm can produce large errors on the quantity itself. The estimated parameters *A* and *B* differ a little according to the sample used. In the French case, let us show the difference in the daily forecast of infected people between the estimation when we use respectively the 28 first samples (Figure-3a) and the 28 last samples (Figure-3b).

**Figure 3:**
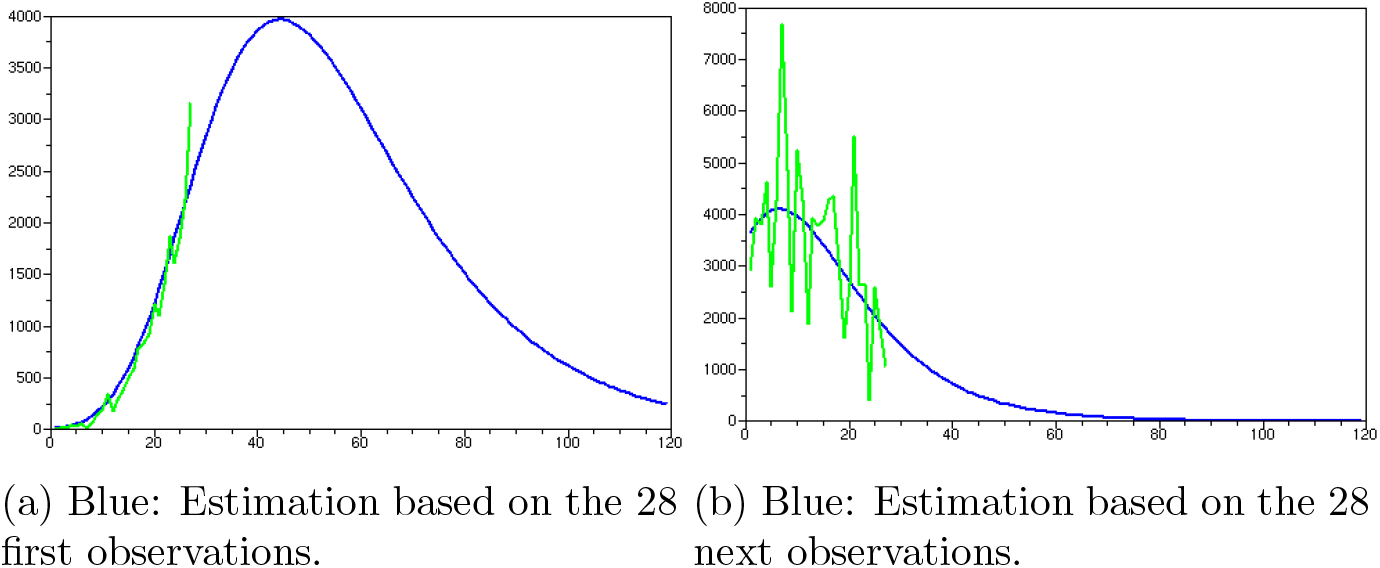
Green: Observed daily infected people in France.

The parameters (*A, B, T*) are equal respectively to (0.050, 0.617, 5) and (0.079, 0.944, 24). Considering the changes of policy in France, we find that the result is quite robust. The forecast of the total number of infected people at the end of the epidemic is given by exp *B/A* that is in the two cases 214 000 and 145 000 respectively.

It is important to notice that the French containment policy has been decided March 17 which is day 22 on Figure-3a and Figure-4. On this date, it was decided to change the covid test policy. France entered in what is called phase^2^ 3 which is not to try to detect anymore all infected people but only the severe cases.

**Figure 4:**
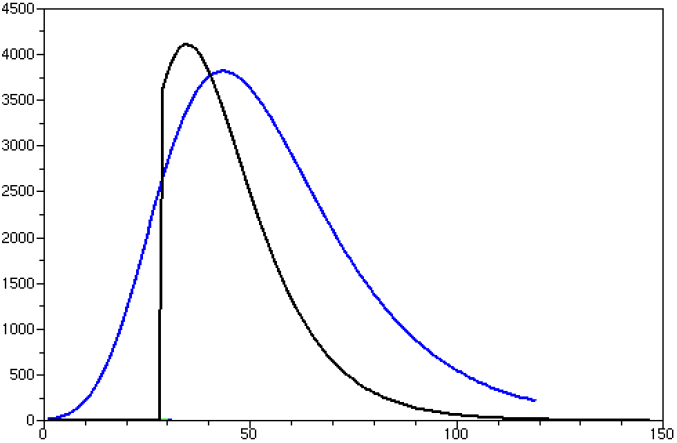
Blue: Daily infected people estimation based on the 28 first observations. Black: estimation based on the 31 last observations.

In Figure-4 we also clearly see the forecast difference and the impact of containment policy. The containment is not as crucial as it is often said.

Let us give the prediction and observation of the daily infected-recovered people obtained with this simple model for two other countries: Germany and Italy.

## 4 Analysis of French containment policy based on these epidemic estimations

Let us discuss if the French decision of containment taken on the 17th of March 2020 was justified. This is why we have to estimate the number of deaths avoided by this decision. From this discussion above, we can first determine the avoided number of infected-recovered people *O*. It can be obtained easily by *O* = exp(*B/A*) *−* exp(*B*′*/A*′). On the one hand (*B, A*) are parameters provided before the containment, and on the other hand, (*B*′, *A*′) are parameters at the present time based on observations since the containment decision time has decided. It is approximately equal to *O* = exp(0.617*/*0.05) *−* exp(0.943*/*0.079) = 76 000.

Now, to give an estimate of avoided death number, we have to use a death rate defined as the number of death in hospitals divided by the number of observed infected-recovered people counted in hospitals. But this death rate changes a lot over time (see Figure-6). We can see the rate is approximately constant and equals to 0.02 during phase 1 and phase 2 of the French covid policy where active research of infected people was done. During phase 3, only severe covid cases were detected and the death rate is increasing almost linearly. If we take the final death rate of 0.1, we obtain 7 600 avoided deaths.

**Figure 5:**
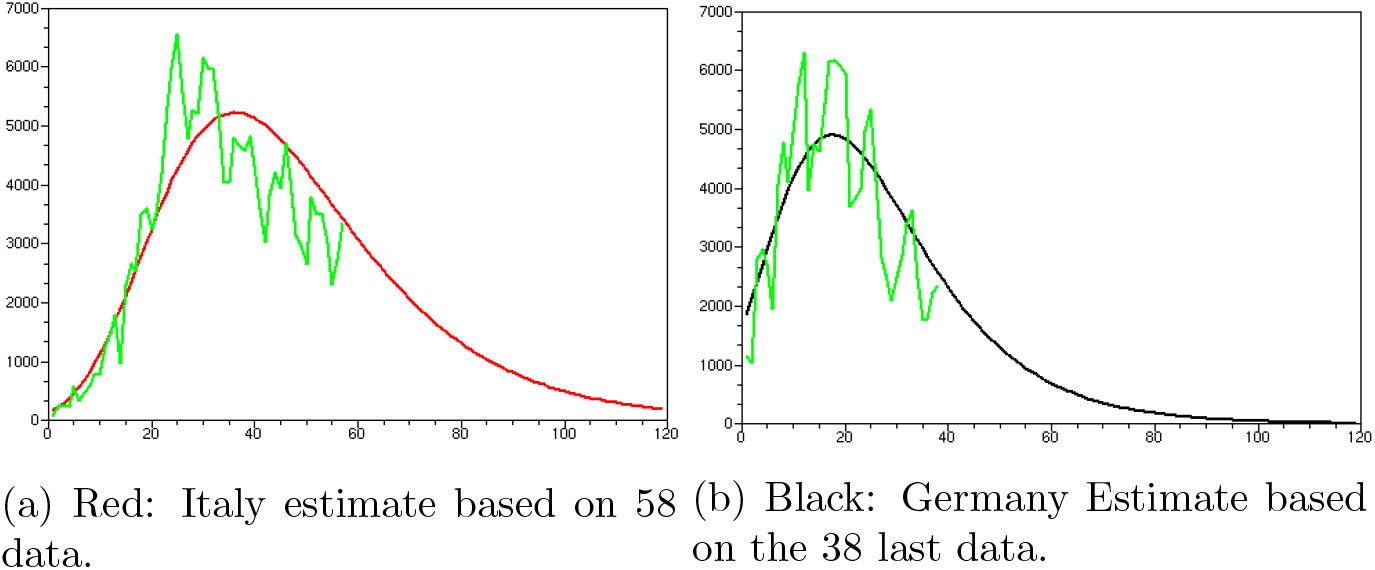
Green: Observed daily infected-recovered people in Italy and Germany (last data 22 April).

**Figure 6:**
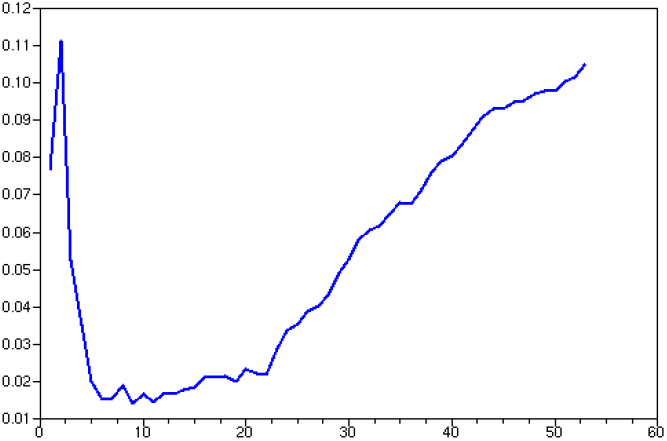
Covid Death Rate in France as function of time.

Pr. Raoult’s study obtains a death rate of 0.004 at the date of April 23, 2020, from 3040 patients having followed his care [8]. Because Pr. Raoult was very active in testing patients, this rate must be compared with the French rate before phase 3 which is 0.02, five times the rate obtained by Pr. Raoult. What would have happened if the French government had followed his advice “test-treat-no confine” instead of following the advice of the scientific committee? The human cost would have been 1 500 deaths for the absence of containment. But the 12 000 deaths in hospitals would have been divided by 5 that is 9 500 deaths avoided. At the end, follow-up on the advice of Pr. Raould would have saved 8 000 of hospital deaths.

## 5 Conclusion

SIR models are not effective to estimate covid epidemic. An alternative way to obtain a correct prediction is to fit a linear time-invariant first-order dynamical model on the logarithm of the number of observed infected-recovered people. Based on this predictor and the efficiency of the Pr. Raoult’s treatment we can assert with a reasonable degree of confidence, that the “contain and wait for a proved treatment” policy decided by the French government is an error expensive in human lives.

## Data Availability

wikipedia

## Footnotes

1 in fact the *I* variable is the contagious people not the infected people which is rather *T*.

2 The three phases of the French covid policy were: – in phase 1 we try to find the first positive case, – in phase 2 we no longer try to find anymore the initial case but all the positive ones, – in phase 3 we no longer try to find any more all the positive cases but only the severe ones that require hospitalization.

## Notes

### Competing Interest Statement

The authors have declared no competing interest.

### Funding Statement

No funding has been used

## References

[1] “Neil Ferguson”, L’Obs nbr. 2892, p. 11, 9-15 april 2020.

[2] R. Bhardwaj “A Predictive Model for the Evolution of COVID-19”, medRxiv preprint, doi.org/10.1101/2020.04.13.20063271.

[3] Imperial College Covid-19 Response Team, “Estimating the number of infections and the impact of non-pharmaceutical interventions on COVID-19 in 11 European countries”, doi.org/10.25561/77731.

[4] Wikipedia: “Mathematical modelling of infectious disease”.

[5] W.O. Kermack, A.G. McKendrick, “A Contribution to the Mathematical Theory of Epidemics”. Proceedings of the Royal Society A: Mathematical, Physical and Engineering Sciences. 115 (772): 700, 1927. doi:10.1098/rspa.1927.0118. JSTOR 94815.

[6] Wikipedia: “Reed–Frost model”, 1928.

[7] Gautret et al., “Hydroxychloroquine and azithromycin as a treatment of COVID-19: results of an open-label non-randomized clinical trial.” International Journal of Antimicrobial Agents – In Press 17 March 2020– DOI : 10.1016/j.ijantimicag.2020.105949

[8] https://www.mediterranee-infection.com/covid-19/.

[9] https://fr.wikipedia.org/wiki/Pandémie_de_Covid-19en_France.

[10] https://fr.wikipedia.org/wiki/Pandémie_de_Covid-19en_Italie.

[11] https://fr.wikipedia.org/wiki/Pandémie_de_Covid-19_en_Allemagne.

